# The Novel Psychoactive Substances Epidemic: a Scientometric Perspective

**DOI:** 10.1101/2022.10.16.22281132

**Authors:** Michelle Jin Yee Neoh, Alessandro Carollo, Mengyu Lim, Ornella Corazza, Aurora Coppola, Gianluca Esposito

## Abstract

The unprecedented proliferation of novel psychoactive substances (NPS) in the illicit drug market has been a public health concern since their emergence in the 2000s. Their consumption can pose a severe health risks as their mechanism of action is poorly understood and their level of toxicity is high mainly due to the diffusion of very potent synthetic cannabinoids and synthetic opioids. This study systemically analyses the evolution of the scientific literature on NPS to gain a better understanding of the areas of major research interests and how they interlink. Findings indicate that the published evidence covers clusters focused on classes of NPS that have received widespread media attention, such as mephedrone and fentanyl, and have largely been concerned with the pharmacological and the toxicological profiles of these substances. This scientometric perspective also provides greater insight into the knowledge gaps within this new and rapidly growing field of study and highlights the need for an interdisciplinary approach in tackling the NPS epidemic.

## 1 Introduction

In recent years, there has been an unprecedented proliferation of novel psychoactive substances (NPS) in the drug market globally with 1124 substances being reported to the United Nations Office on Drugs and Crime by December 2021 [1]. The term NPS is typically used to refer to substances originally designed as legal alternatives to controlled substances, such as analogues of existing controlled drugs and pharmaceutical products designed to mimic their actions and effects [2, 3]. The emergence of NPS in the illicit drug market poses a serious challenge for drug policy and public health. The ease and speed with which NPS are continuously developed and reintroduced in the market as analogues or derivatives with small chemical changes in order to circumvent drug enforcement and legislation makes these substances difficult to legally regulate and control. In addition, the potency, pharmacological effects and risk profiles of NPS are diverse and generally unknown, where users are often misinformed and lack awareness of the contents of the NPS they are consuming. The properties and long-term effects of NPS are not well understood and few studies examining most of these new substances have been published [4].

Adverse health effects including suicide and fatalities have been documented in association with and attributed to NPS use [5] and the most frequent classes of NPS are associated with medical risks [6]. In an analysis of hospital emergency data by the European Drug Emergencies Network, 9% of all drug-related emergencies involved NPS [7]. Reports have also cited correlations between NPS use and increased spread of diseases such as HIV and hepatitis C [8]. Although case reports and case series form the bulk of research informing our understanding of NPS, it is apparent that there are significant health risks associated with NPS use.

Although the prevalence of NPS in the general population appears to be low, their consumption among younger age groups is higher and warrants concern. A systematic review found that most studies showed 3% or less of adult populations reported recent use of NPS, typically in the past year, compared to an estimated 1 in 10 young people [9]. Similarly, an estimated 1-8% of school students have used NPS at some point [10], indicating a worrying trend of drug use among youth. However, it is important to note that such self-report data on NPS use are most likely not an accurate representation of actual market penetration and user uptake of NPS due to the sheer speed in which NPS are introduced to market as well as unwitting consumers.

The aim of this study is to identify the most relevant publications, the nature of their inquiry and associate literature gaps in the field of NPS. We conducted a document co-citation analysis (DCA) [11, 12] to analyse references and the relevance of these publications in the existing literature on NPS. By clustering publications according to common research domains, articles with significant contributions to the literature can be identified while highlighting the coverage and gaps in existing literature. The scientometric results can illuminate research and identifying publication trends, the links between different works and scientific fields involved in the exploration of issues relating to NPS. The scientometric approach has been employed in neuroscience [13] and digital psychology [14].

## 2 Material and Methods

Publications were downloaded from Scopus in accordance with the standard and established scientometric procedures [11]. We used the following search string TITLE-ABS-KEY(“novel psychoactive substanc*” OR “new psychoactive substanc*”) AND (LIMIT-TO (LANGUAGE, “English”)) and found a total of 2,365 documents published from 1 January 2010 to 13 June 2022. By limiting the search to publications in English, the analysis will be built on international scientific literature in the field, allowing for a more standardised and rigorous examination of existing work [15].

### 2.1 Data import on CiteSpace

Scientometric analysis was conducted using CiteSpace software (Version 6.1.R2) and the articles downloaded from Scopus were imported into the software. 106,911 of a total of 111,855 references (95.58%) cited by the 2,365 articles were valid (Figure 1). A “valid” reference is one that contains seven key pieces of information: author, year of publication, title, source, volume, pages and DOI [11]. Due to irregularities in the citation format, a number of references were considered invalid. Negligible losses in references (1-5%) commonly occur during the data import to the CiteSpace software [16]. The CiteSpace function (Remove Alias) was turned ON to eliminate repeated or identical entries.

**Figure 1:**
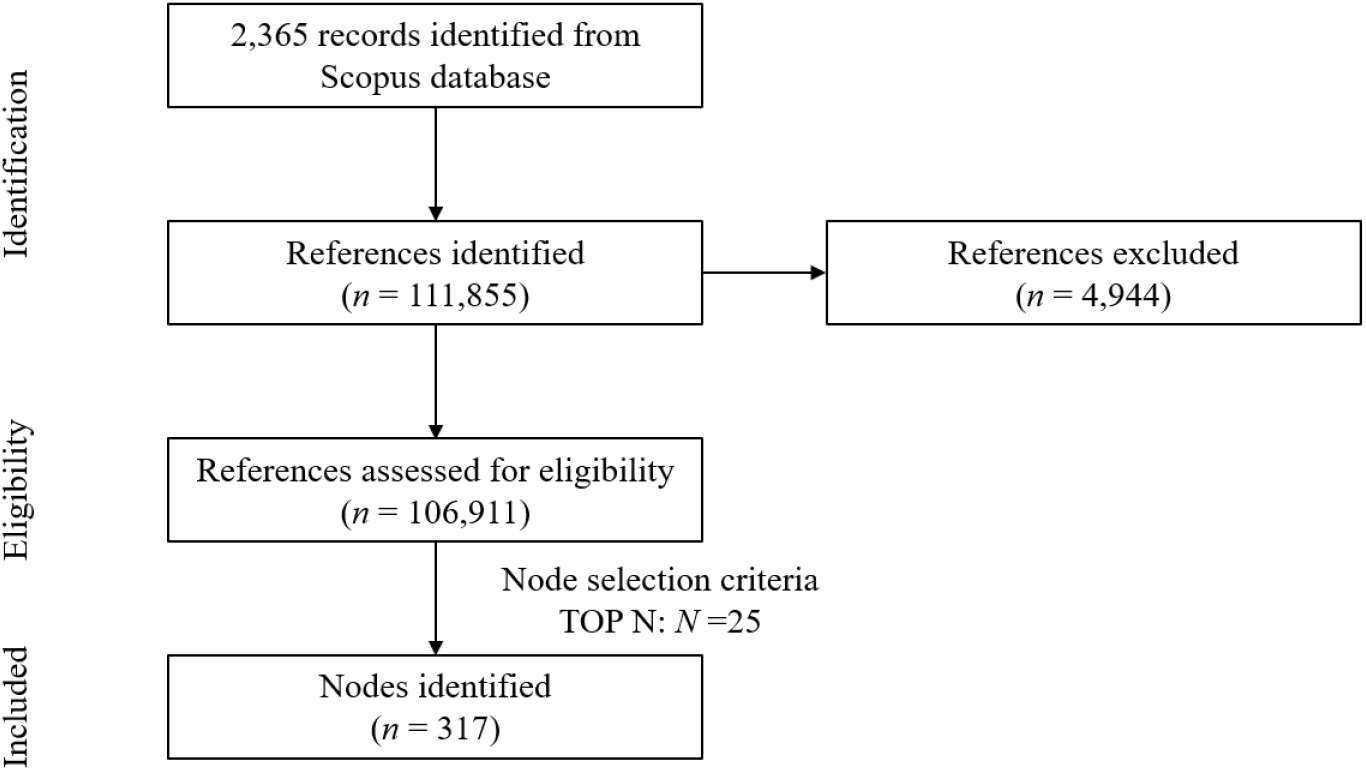
PRISMA flowchart for search criteria and reference eligibility

### 2.2 Document Co-Citation Analysis (DCA) and Optimisation of Parameters

DCA was conducted to determine the main research domains in NPS literature. DCA is based on the frequency with which two or more papers are cited together in source articles [17]. The assumption is that frequent co-citations among articles reflect common research trends and intellectual domains in the literature [12, 18]. The network resulting from the DCA is composed of documents frequently cited together along with the documents that cite them (i.e. articles downloaded from Scopus).

DCA parameters were optimised in order to obtain a balanced network of documents. Several DCAs were computed and compared, each with a different setting for one of three node selection criteria; g-index, TOP *N*, TOP *N* %, as done in [13, 15, 18, 19, 14, 20]. The node selection criteria are *a priori* settings that define the criterion used for selecting articles to be included in the network, and consequently, determine the final network of articles being generated. The g-index is a measure of the citation scores of an author’s top publications [21, 22]. It represents the largest number that equals the average number of citations of the most highly cited g publications [16]. TOP *N* and TOP *N* % are criteria used to select *N* and *N* % most cited within a time slice (1 year was used in this study) as network nodes respectively [11].

In order to generate the final optimal network, node selection criteria were varied together with their scale factor values, which refer to the chosen numeric values used as thresholds for the respective node selection criteria [14]. Specifically, DCAs with the following node selection criteria were compared: g-index with scale factor *k* set at 10, 15, 25, 50, TOP *N* with scale factor *N* set at 25, 50 and TOP *N* % with scale factor *N* set at 10. To determine the node selection criteria and scale factor to use for the generation of the final network, the overall effects on the structural metrics of the generated network, the number of nodes included and clusters identified were compared. After comparison of these metrics, TOP *N* with *N* at 25 was the parameter which was used to generate the final network of articles.

### 2.3. Metrics

Structural and temporal metrics are used in describing CiteSpace results. Structural metrics consist of (i) *modularity-Q*, (ii) *silhouette scores* and (iii) *betweenness centrality*. Modularity-Q values range from 0 to 1 and indicates the degree to which the network can be decomposed into single groups of nodes, which are referred to as modules or clusters [23]. High modularity-Q values indicate a well-structured network [12]. Silhouette scores are a measure of inner consistency - cohesion and separation - of the modules [24]. Silhouette scores vary from -1 to +1, where higher values represent high separation from other modules and internal consistency [25]. Betweenness centrality represents the degree to which a node connects an arbitrary pair of nodes in the network [11, 26]. Betweenness centrality values range from 0 to 1, where groundbreaking and revolutionary works in the scientific literature typically score higher [27].

Temporal metrics consist of (i) *citation burstness* and (ii) *sigma*. The Kleinberg’s algorithm [28] is used to calculate citation burstness, which indicates a sudden increase in the number of citations of an article in a given time frame [29]. Sigma is calculated with the equation (centrality + 1)^burstness^ and indicates the novelty of a document and influence on the overall network [30].

The overall configuration of the generated network and identified clusters of references were examined with modularity-Q and silhouette scores. The attributes of single nodes in the network were examined using betweenness centrality and the temporal metrics.

## 3 Results

### 3.1 Structural metrics

The final optimised network obtained from the DCA consisted of 317 nodes and 1,327 links, indicating an average of 4.15 connections with other references for each node (Figure 2). The network had a modularity-Q index of 0.6181 and a mean silhouette score of 0.8445, indicating medium divisibility of the network into clusters that are highly homogenous.

**Figure 2:**
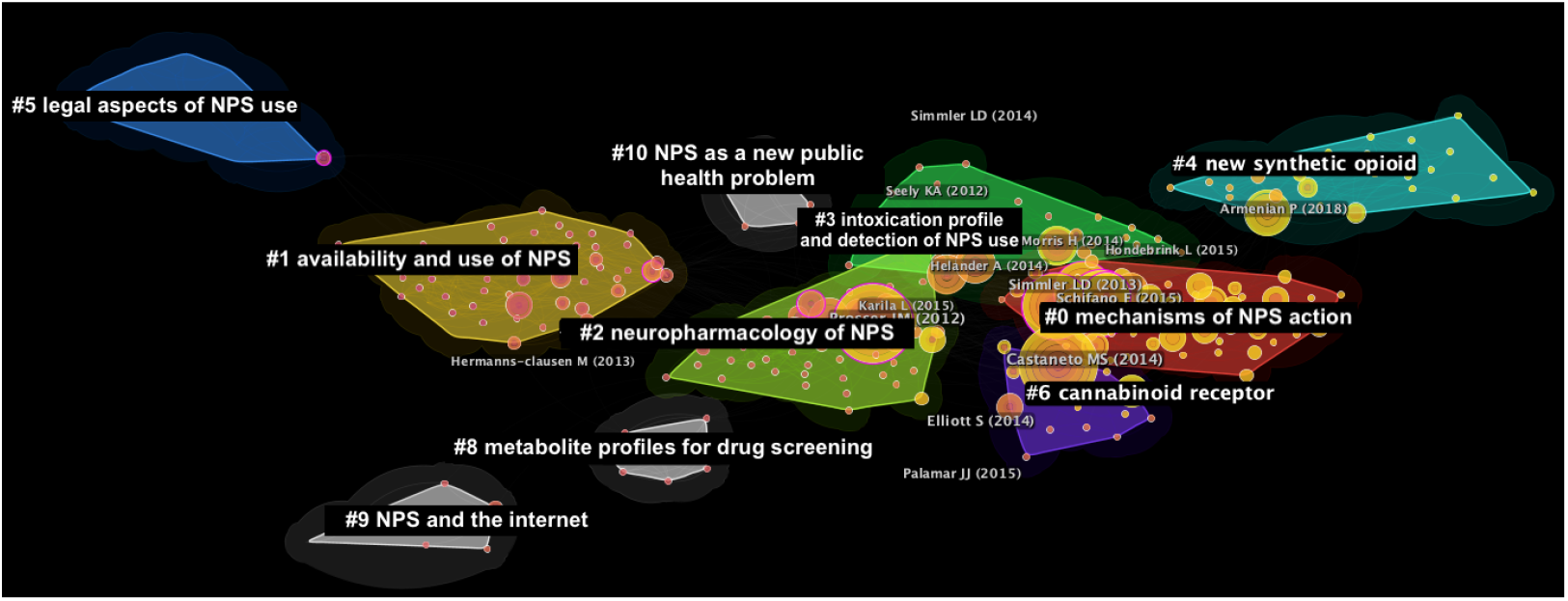
Network of publications generated through the document co-citation analysis (DCA). The major clusters are grouped by colour.

### 3.2. Citation burstness

A total of 64 documents were found to exhibit a citation burstness (Table 1). 20 of these documents belong to cluster #0, 13 to cluster #1, 16 to cluster #2, 4 to cluster #3, 3 to cluster #4, 1 to cluster #5, 4 to cluster #6 and 1 to cluster #9. The article with the highest burst strength was authored by Hondebrink et al. in 2015 [31] with a score of 12.43. The burst began in 2017 and ended in 2019. The article with the longest burst duration was authored by Castaneto et al. in 2014 [32] with a burst duration of 5 years from 2017 to 2022. The article with the highest sigma value was authored by Hermanns-clausen in 2013 [33] with a sigma value of 2.44.

**Table 1:** Top 15 publications in terms of burst strength.

| Reference | Burst | Year | Start of burstness | End of burstness | Burst duration | Centrality | Sigma |
| --- | --- | --- | --- | --- | --- | --- | --- |
| Hondebrink et al. [31] | 12.43 | 2015 | 2017 | 2019 | 2 | 0.01 | 1.10 |
| Armenian et al. [34] | 11.68 | 2018 | 2019 | 2022 | 3 | 0.02 | 1.19 |
| Peacock et al. [9] | 10.97 | 2019 | 2020 | 2022 | 2 | 0.01 | 1.08 |
| Brandt et al. [35] | 10.93 | 2014 | 2016 | 2017 | 1 | 0.00 | 1.00 |
| Rickli et al. [36] | 10.58 | 2015 | 2017 | 2018 | 1 | 0.00 | 1.04 |
| Morris et al. [37] | 10.07 | 2014 | 2017 | 2018 | 1 | 0.05 | 1.67 |
| Measham et al. [38] | 9.46 | 2011 | 2012 | 2014 | 2 | 0.07 | 1.90 |
| Hermanns-clausen et al. [33] | 9.44 | 2013 | 2014 | 2017 | 3 | 0.01 | 2.44 |
| Kraemer et al. [39] | 8.62 | 2019 | 2020 | 2022 | 2 | 0.02 | 1.17 |
| Seely et al. [40] | 8.51 | 2012 | 2015 | 2018 | 3 | 0.04 | 1.41 |
| Richter et al. [41] | 8.26 | 2017 | 2019 | 2020 | 1 | 0.00 | 1.01 |
| Baumann et al. [42] | 7.92 | 2012 | 2016 | 2017 | 1 | 0.01 | 1.09 |
| Prosser et al. [43] | 7.81 | 2012 | 2016 | 2017 | 1 | 0.17 | 3.50 |
| Majchrzak et al. [44] | 7.68 | 2018 | 2020 | 2022 | 2 | 0.01 | 1.12 |
| Favretto et al. [45] | 7.55 | 2013 | 2015 | 2017 | 2 | 0.01 | 1.10 |

### 3.3. Thematic clusters

Ten major clusters were identified (Figure 2, Table 2). The largest cluster #0 consisted of 59 nodes and had a silhouette score of 0.716, where the constituent references were published in 2019 on average. The cluster was labelled “Mechanisms of NPS action”. Second, cluster #1 consisted of 56 nodes and had a silhouette score of 0.84, where the constituent references were published in 2012 on average. The cluster was labelled “Availability and use of NPS”. Third, cluster #2 consisted of 49 nodes and had a silhouette score of 0.809, where the constituent references were published in 2014 on average. The cluster was labelled “Neuropharmacology of NPS”.

**Table 2:**
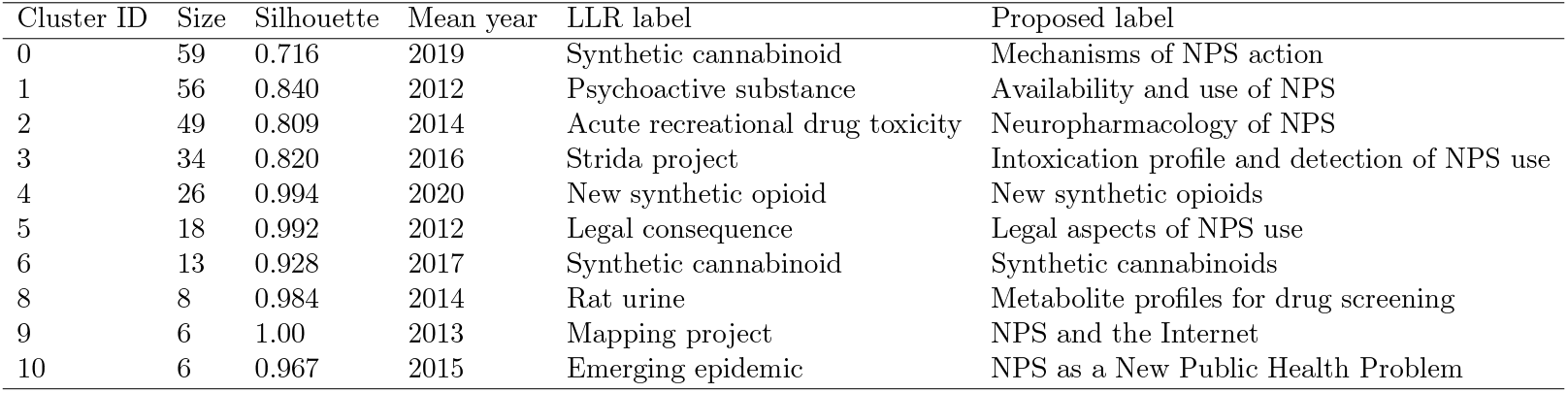
Metrics of the 10 clusters identified with the DCA. Log-likelihood Ratio (LLR) labels are automatically generated by the software.

## 4 Discussion

In this section, we will discuss each cluster in greater detail in chronological order, by the average year of publication of the cluster. Each cluster will be analysed in terms of both the citing articles and the cited references. The main citing articles for each cluster will be highlighted, together with its coverage and Global Citing Score (GCS). Coverage refers to the number of articles in the cluster that were cited by the citing article and GCS refers to the total number of citations received by a paper as indexed on Scopus.

### 4.1. Cluster #5: Legal aspects of NPS

The major citing articles in Cluster #5 were authored by Bilinski et al. [46] with a coverage of 14 articles and GCS of 6, and B McNabb et al. [47] with a coverage of 11 articles and GCS of 11. Cluster #5 appears to be one of the earlier groups of work investigating NPS emerging on the drug market, their effects and their legal status, as seen from the mean year of publication (2012) and a significant number of cited references conducted in animal models prior to 2010 [48, 49, 50, 51, 52]. The primary theme of this cluster appears to be legal concerns of NPS use following their entrance into the market, where the citing articles focused on the legality of designer drugs touted as “legal” in the drug market [46, 47]. Accordingly, there were also cited references looking into the legal aspects of NPS use [53, 54].

### 4.2. Cluster #1: Availability and use of NPS

The major citing articles in Cluster #1 were authored by Smith and Garlich [55] with a coverage of 21 articles and GCS of 14, Wood and Dargan [56] with a coverage of 16 articles and GCS of 7, and Wood and Dargan [57] with a coverage of 14 articles and GCS of 54. Cluster #1 appears to be a body of work that emerged in response to the series of bans on mephedrone as well as other cathinones across multiple countries across the world in 2010, as evidenced by the mean year of publication (2012). The focus of both citing articles and the cited references appeared to be the same, which primarily revolved about the effects, prevalence and use of mephedrone and other cathinones. A significant number of cited references examined their use [38, 58, 59] and continued presence in the composition of existing drug products available in the market [60, 35] despite their legal status, as well as their effects (e.g. [61, 59, 62, 63]). This was also the case for many of the citing articles which also looked at the effects of mephedrone and other cathinones [57, 64, 56, 65].

### 4.3. Cluster #9: NPS and the Internet

The major citing articles in Cluster #9 were authored by Deluca et al. [66] with a coverage of 3 articles and GCS of 139, Cinosi et al. [67] with a coverage of 2 articles and GCS of 99, and Corazza et al. [68] with a coverage of 2 articles and GCS of 52. The main theme of Cluster #9 is the role of the Internet as a source for information on NPS as well as a virtual marketplace for NPS. It is clear from the cited articles that the availability of NPS through online channels is a key concern, especially since consumer reach and engagement on the Internet is unrivalled in comparison to a brick-and-mortar store, as well as the information on NPS available on the Internet (e.g. [69, 70, 71, 72]). Targeting the Internet through researching and designing web-based interventions such as web monitoring and information dissemination is thus a recommendation and/or central focus of the citing articles in Cluster #9 (e.g. [66, 68]). Research in Cluster #9 highlights the information war on the Internet as a key aspect of the NPS crisis and the importance of developing information and communication technology interventions.

### 4.4. Cluster #8: Metabolite profiles for drug screening

The two major citing articles in Cluster #8 were authored by Welter et al. [73] with a coverage of 8 articles and GCS of 25 and Welter et al. [74] with a coverage of 72 articles and GCS of 6. Both of these citing articles investigated methods for detecting of designer drugs through screening the metabolic products present in rat urine by applying gas chromatography-mass spectroscopy and/or liquid chromatography methods. This strongly suggests the cluster is geared towards the application of these chemical methods to NPS detection. Majority of the cited articles also report the results of applying these methods to detect drug metabolites in urine (e.g. [75, 76, 77, 78, 79]).

### 4.5. Cluster #2: Neuropharmacology of NPS

The major citing articles in Cluster #2 were authored by Zawilska and Andrzejczak [80] with a coverage of 10 articles and GCS of 120, Papaseit et al. [81] with a coverage of 10 articles and GCS of 20, and [82] with a coverage of 10 articles and GCS of 88, which were all reviews on the pharmacology of NPS use. The cluster is centred in elucidating the pharmacology of the various classes of NPS, which the citing articles covered from cathinones (e.g. [83, 84]), cannabinoids (e.g. [85]), hallucinogens [86] and stimulants [87]. The same theme is reflected in the cited articles, which were mostly concerned with the toxicological findings from NPS use [33, 88, 43, 89, 90]. Hence, Cluster #2 appears to be a body of work relating pharmacological properties of NPS to their toxicity and risk of fatality.

### 4.6. Cluster #10: NPS as a New Public Health Problem

The major citing articles in Cluster #10 were authored by Zawilska and Andrzejczak [80] with a coverage of 4 articles and GCS of 36, Karila et al. [91] with a coverage of 3 articles and GCS of 129, and Rickli et al. [36] with a coverage of 3 articles and GCS of 129. The focus of these reviews aims to consolidate information on properties of NPS in relation to their propensity as a public health threat by presenting clinical data. The findings in the cited references also presents clinical data [92], including psychosis [93].

### 4.7. Cluster #3: Intoxication profile of NPS use

The major citing articles in Cluster #3 were authored by Schifano et al. [94] with a coverage of 7 articles and GCS of 178, Salomone [95] with a coverage of 7 articles and GCS of 85, and Castaneto et al. [96] with a coverage of 7 articles and GCS of 4, which were all reviews on the pharmacology of NPS use. The cluster is composed of a group of work collecting data on the intoxication profiles of NPS users (e.g. [97, 98, 99, 100, 101]). Notably, a significant number of the citing articles analysed data from the STRIDA project in Sweden, which monitored occurrences and health hazards of NPS and includes information from about 2,600 cases of suspected NPS intoxications across the period of 2010-2016 [102, 103, 104, 105, 106, 107]. Similarly, the cited references were also research on intoxication profiles and fatality from NPS use (e.g. [108, 109, 110, 111, 112, 113, 114, 115]). The size of the cluster alludes to the public health threat that NPS use poses, as well as the importance of the field of emergency medicine in monitoring adverse events relating to drug use which can provide much needed data on the toxicological and risk profile of NPS use.

### 4.8. Cluster #6: Synthetic cannabinoids

The major citing articles in Cluster #6 were authored by Cannaert et al. [116] with a coverage of 5 articles and GCS of 22, Ametovski et al. [117] with a coverage of 5 articles and GCS of 5, and [118] with a coverage of 4 articles and GCS of 18. Considering that marijuana - which contains cannabis - is one of the most commonly use drugs in the world [119], it is not surprising that synthetic cannabinoids gained popularity as a “legal” alternative in countries - “Spice” being an example of a brand name cannabinoid drug - where cannabis is controlled. Hence, this cluster of work examining the effects of the recreational use of these synthetic cannabinoids appears to be in response to their widespread use and market penetration in the recreational drug market, especially considering its mean year of publication in 2017, which was around the time period when cannabis began to be legalised for recreational use and sale as a consumer product across a number of countries such as Canada and the USA. The citing articles include research into the pharamcology [120, 121, 122, 123], toxicology [124, 125, 126, 127] as well as a number of articles pointing towards the sheer volume and continued emergence of variations of synthetic cannabinoids [118, 128, 129]. Similarly, a bulk of the cited references also focus mainly on synthetic cannabinoids and their fatalities associated with their use [32, 130, 131, 132].

### 4.9. Cluster #0: Mechanisms of NPS action

The major citing articles in Cluster #0 were authored by Miliano et al. [82] with a coverage of 13 articles and GCS of 88, Rudin et al. [133] with a coverage of 12 articles and GCS of 9, and Ellefsen et al. [83] with a coverage of 11 articles and GCS of 45, which were all reviews on the pharmacological effects of NPS use. Firstly, many of the cited references in the cluster discussed fatalities from NPS use (e.g. [31, 39, 134, 135]), indicating that research on NPS is driven towards characterising the risk profile of NPS use in terms of clinical outcomes and fatalities. Secondly, each of the major citing articles were focused on different classes of NPS; Miliano et al. [82] focused on cannabimimetics and amphetamine stimulants, Rudin et al. [133] on stimulant and psychedelic NPS and Ellefsen et al. [83] on synthetic cathinones. This highlights the diversity of NPS classes available, which poses an immense challenge to the scientific community as evidenced by the constant refrain of a lack of understanding of these mechanisms in many citing articles. A tremendous effort is required in organising and conducting research into understanding the drug action and effects of the different classes of NPS, as seen from the size of this cluster. A defining characteristic of the NPS phenomenon, the rapid development of NPS entering the drug market suggests the continual need for research to update the scientific understanding of the mechanisms of drug action, which is suggested by the relatively recent mean year of publication (2019). Many of the references being cited were also investigating the pharmacological actions and toxicity of NPS (e.g. [36, 41, 136, 137, 138, 139]), further highlighting the need to constantly build onto existing scientific knowledge because of the sheer speed of NPS variations being introduced in the market.

### 4.10. Cluster #4: New synthetic opioids

The major citing articles in Cluster #4 were authored by Vandeputte et al. [140] with a coverage of 7 articles and GCS of 1, Vandeputte et al. [141] with a coverage of 7 articles and GCS of 3, and [142] with a coverage of 7 articles and GCS of 10. Given its recent mean year of publication (2020), the cluster is very likely to be a group of work in response to rising use of synthetic opioids globally. While the opioid crisis is not a new phenomenon, a spotlight has been thrown onto synthetic opioid use in the wake of the recent surge in opioid overdose deaths in the US - an opioid overdose epidemic driven by synthetic opioids, fentanyl and its derivatives in particular [143, 144, 145]. The cited reference with the highest burst strengths in this cluster focuses on fentanyl and other novel synthetic opioids [34, 146, 147], supporting this notion. Many citing articles were concerned with opioid-related deaths [39, 148, 149, 150] and a significant number of articles pointed towards the rapidly surging numbers globally and its implications on public health [151, 152, 153].

### 4.11. Limitations

There are a number of limitations of the scientometric approach. The DCA is based on the quantity of citations and co-citation patterns in the retrieved references and does not provide a qualitative perspective for these citation patterns per se. Hence, the rationale for the cited references are not immediately clear from the scientometric analysis.

## 5 Conclusions

The NPS phenomenon is a major cause for concern in countries across the globe and is a war on drugs fought on many fronts, as evidenced by the diversity of scientific fields represented in the network of articles derived from the DCA. Given the challenges that NPS pose in terms of drug monitoring, surveillance, control and public health responses, understanding the scientific literature and disciplines involved in research on NPS is a valuable contribution to the field which can be harnessed in supporting drug policy and public health response. Such an analysis can highlight the research gaps in NPS to direct attention to and facilitating interdisciplinary research based on the research domains and links identified. In addition, producers of NPS often mine knowledge from scientific journals and an understanding of the information that is available can be useful in targeting and informing drug policy. Thus, drug policy, monitoring and control can benefit from coordinated, interdisciplinary efforts to inform and develop a multi-pronged approach to the NPS crisis in order to safeguard public health.

## Data Availability

All data produced in the present study are available upon reasonable request to the authors

## Conflict of interests

The authors declare no conflict of interest.

## Notes

### Competing Interest Statement

The authors have declared no competing interest.

### Funding Statement

This study did not receive any funding

